## Supplementary file for "Three components of glucose dynamics – mean, variance, and autocorrelation – are independently associated with coronary plaque vulnerability"

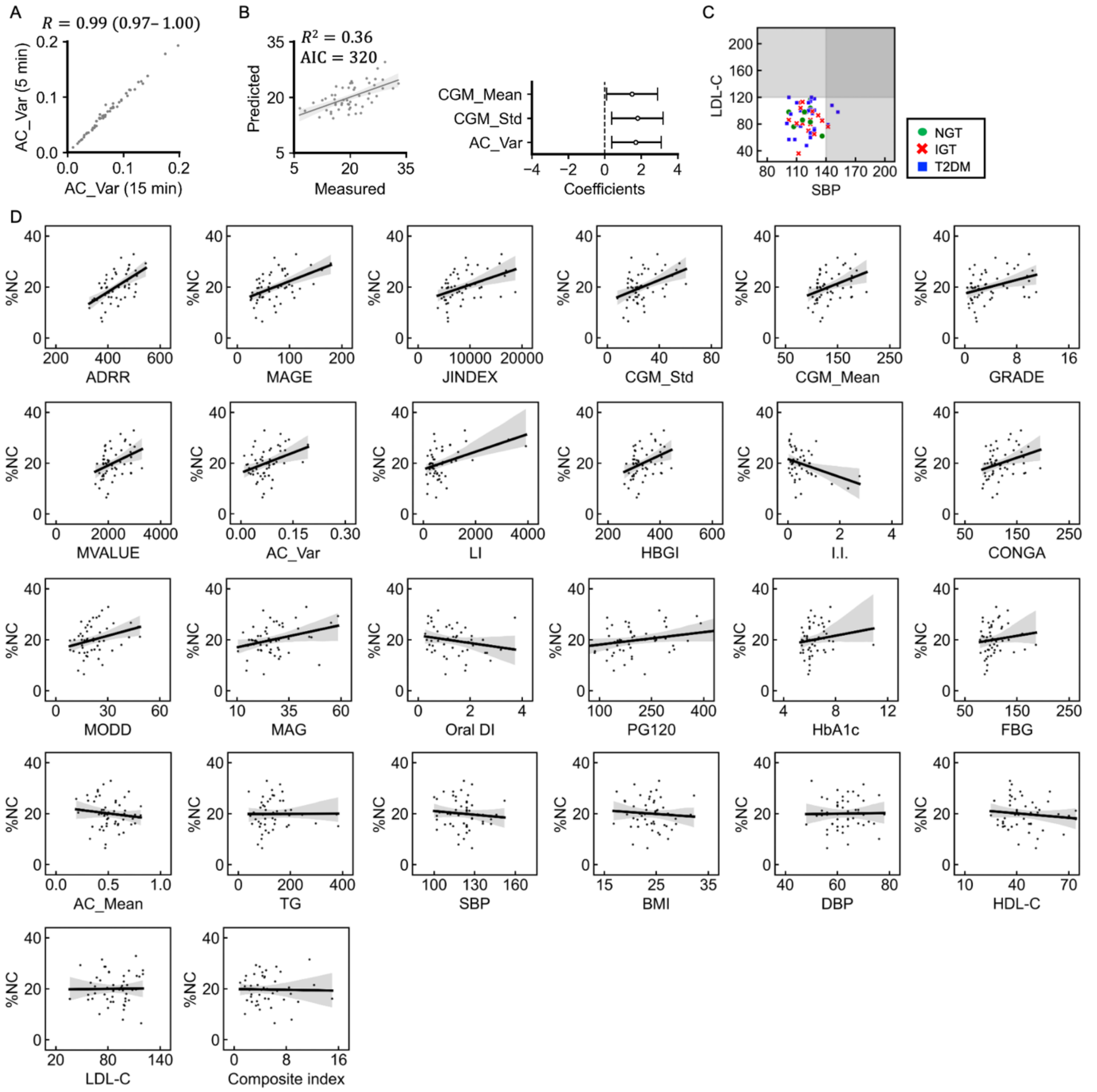


**Figure S1. Relationship among clinical parameters.**

(A) Scatter plot of the AC_Var calculated from 15 min-intervals and that calculated from 5 min-intervals. R is Spearman's correlation coefficient, and the value in parentheses is the 95% CI.

(B) Multiple regression analysis between %NC and CGM_Mean, CGM_Std, and AC_Var. These CGM-derived indices were calculated from CGM data of 15 min-intervals. Scatter plots for predicted %NC versus measured %NC (the left). Each point corresponds to the values for a single individual. Bars represent the 95% CIs of the coefficients of the regression models (the right).

(C) Scatter plot for SBP and LDL-C. Gray shaded areas indicate the range of values for high SBP (>140 mmHg) or high LDL-C (>120 mg/dL). Each point corresponds to the values for a single participant. Green circles, red cross marks, and blue squares indicate NGT, IGT, and T2DM, respectively.

(D) Scatter plots and fitted linear regression lines for each clinical index versus %NC. Each point corresponds to the values for a single individual. Gray shaded area indicates the 95% CI.

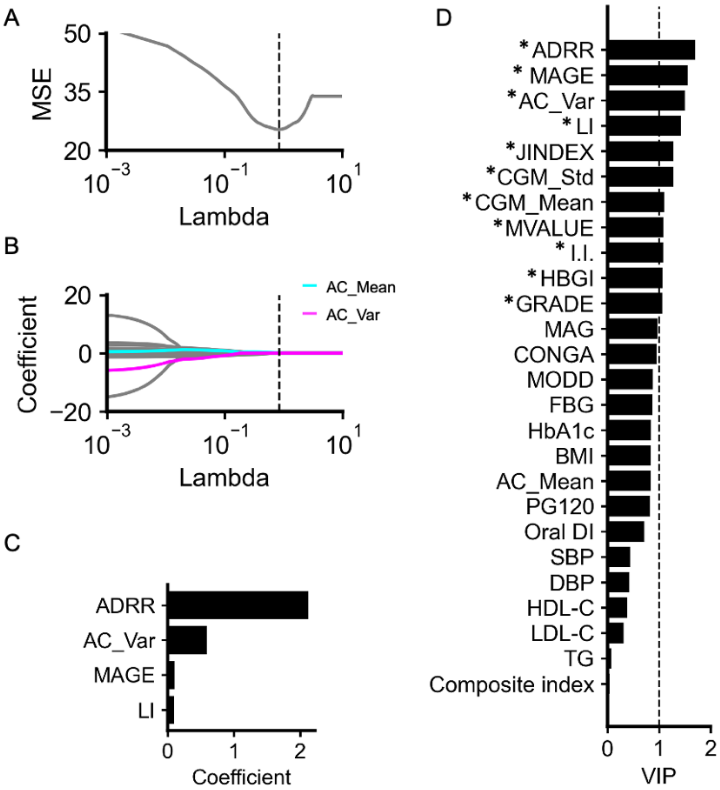


**Figure S2. LASSO and PLS regression analyses for predicting %NC including SBP, DBP, TGs, LDL-C, and HDL-C.**

(A) Relationship between regularization coefficients (lambda) and the MSE based on the leave-one-out cross-validation in predicting %NC. Dotted vertical line indicates the optimal lambda that provides the least MSE. The optimal lambda was 0.849.

(B) LASSO regularization paths along the lambda in predicting %NC. Cyan, magenta, and gray lines indicate the estimated coefficients of AC_Mean, AC_Var, and the other input variables, respectively. Dotted vertical line indicates the optimal lambda.

(C) Estimated coefficients with the optimal lambda. Only variables with non-zero coefficients are shown. Input variables include the following 26 variables: BMI, SBP, DBP, TGs, LDL-C, HDL-C, FBG, HbA1c, PG120, I.I., composite index, oral DI, CGM_Mean, CGM_Std, CONGA, LI, JINDEX, HBGI, GRADE, MODD, MAGE, ADRR, MVALUE, MAG, AC_Mean, and AC_Var.

(D) VIP generated from the PLS regression predicting %NC. Variables with a VIP $\geq 1$ (the dotted line) were considered to significantly contribute to the prediction

A

| | 1, Mean | 2, Variance | 3, Autocorrelation | 4 | 5 | 6 |
|---|---|---|---|---|---|---|
| BMI | 0.11 | 0.1 | 0.16 | 0.13 | 0.59 | 0.09 |
| FBG | 0.75 | 0 | 0.08 | 0.08 | 0.29 | 0.23 |
| HbA1c | 0.69 | 0.1 | 0.17 | 0.14 | 0.33 | 0.19 |
| PG120 | 0.58 | 0.21 | 0.1 | 0.41 | 0.37 | 0.11 |
| I.I. | -0.41 | -0.1 | 0 | -0.72 | -0.02 | -0.02 |
| Composite index | -0.05 | 0.01 | 0.04 | 0.05 | -0.42 | 0.05 |
| Oral DI | -0.58 | -0.01 | -0.04 | -0.52 | -0.51 | 0.05 |
| CGM_Mean | 0.98 | 0.13 | 0.03 | 0.13 | 0.05 | -0.04 |
| CONGA | 0.98 | 0.02 | 0.12 | 0.17 | 0.03 | 0.02 |
| GRADE | 0.98 | 0.07 | 0.01 | 0.16 | 0.04 | -0.07 |
| MVALUE | 0.98 | 0.1 | 0.02 | 0.15 | 0.04 | -0.06 |
| CGM_Std | 0.95 | 0.14 | 0.06 | 0.06 | 0.12 | -0.05 |
| HBGI | 0.21 | 0.94 | 0.09 | 0.07 | -0.03 | 0.16 |
| MAGE | 0.17 | 0.86 | -0.13 | 0.25 | -0.03 | 0.07 |
| LI | -0.01 | 0.87 | -0.26 | -0.01 | -0.01 | -0.12 |
| MAG | 0.01 | 0.87 | -0.3 | -0.25 | 0.16 | -0.24 |
| MODD | 0.14 | 0.71 | 0.2 | 0.05 | 0.12 | 0.07 |
| JINDEX | 0.89 | 0.44 | 0.07 | 0.09 | 0.04 | 0.06 |
| ADRR | 0.66 | 0.64 | -0.09 | 0.23 | 0.02 | -0.02 |
| AC_Mean | 0.22 | -0.04 | 0.91 | 0.18 | -0.02 | 0.07 |
| AC_Var | -0 | 0.19 | -0.92 | 0.16 | -0.15 | 0.05 |

Factor loading
1
-1

B

| | 1, Mean | 2, Variance | 3, Autocorrelation | 4 | 5 | 6 |
|---|---|---|---|---|---|---|
| SBP | 0.11 | -0.16 | 0.02 | 0.22 | 0.23 | 0.06 |
| DBP | -0.1 | -0.15 | -0.37 | 0.18 | 0.24 | 0.3 |
| TG | 0.02 | -0.01 | 0.09 | 0.46 | 0.02 | 0.42 |
| LDL-C | -0.07 | -0.15 | -0.1 | 0.13 | -0.06 | 0.39 |
| HDL-C | -0.18 | 0.07 | -0.02 | -0.37 | 0.1 | -0.04 |
| BMI | 0.11 | 0.13 | 0.16 | 0.54 | -0.06 | 0.08 |
| FBG | 0.77 | 0.02 | 0.11 | 0.26 | 0.07 | 0.17 |
| HbA1c | 0.7 | 0.11 | 0.19 | 0.3 | -0.02 | 0.12 |
| PG120 | 0.62 | 0.23 | 0.08 | 0.39 | -0.25 | 0.18 |
| I.I. | -0.47 | -0.1 | -0.03 | -0.14 | 0.65 | -0.14 |
| Composite index | -0.05 | -0 | 0.08 | -0.44 | -0.14 | -0.03 |
| Oral DI | -0.62 | -0.02 | -0.01 | -0.61 | 0.38 | -0.09 |
| CGM_Mean | 0.98 | 0.11 | 0.02 | 0.07 | -0.07 | -0.09 |
| CONGA | 0.98 | 0 | 0.12 | 0.06 | -0.08 | -0.03 |
| JINDEX | 0.98 | 0.05 | 0.01 | 0.07 | -0.09 | -0.11 |
| HBGI | 0.98 | 0.08 | 0.01 | 0.07 | -0.08 | -0.1 |
| MVALUE | 0.95 | 0.12 | 0.05 | 0.13 | 0.02 | -0.1 |
| GRADE | 0.24 | 0.95 | 0.12 | -0.06 | -0.03 | 0.06 |
| CGM_Std | 0.2 | 0.86 | -0.1 | -0.06 | -0.27 | 0 |
| MAGE | -0.01 | 0.86 | -0.25 | -0.03 | -0.05 | -0.18 |
| LI | -0.02 | 0.86 | -0.32 | 0.11 | 0.19 | -0.34 |
| MAG | 0.16 | 0.72 | 0.21 | 0.08 | -0.03 | -0.01 |
| MODD | 0.9 | 0.42 | 0.08 | 0.03 | -0.02 | -0.04 |
| ADRR | 0.69 | 0.63 | -0.08 | 0.04 | -0.17 | -0.05 |
| AC_Mean | 0.24 | -0.05 | 0.89 | 0.06 | -0.1 | 0.08 |
| AC_Var | 0.03 | 0.2 | -0.93 | -0.18 | -0.16 | 0.16 |

**Figure S3. Factor analyses of the clinical parameters.**

Factor analyses after orthogonal rotation. The values were based on the factor loadings. The columns represent each factor. The rows represent input indices. The analyses with the 21 variables (A), and the 26 variables (B).

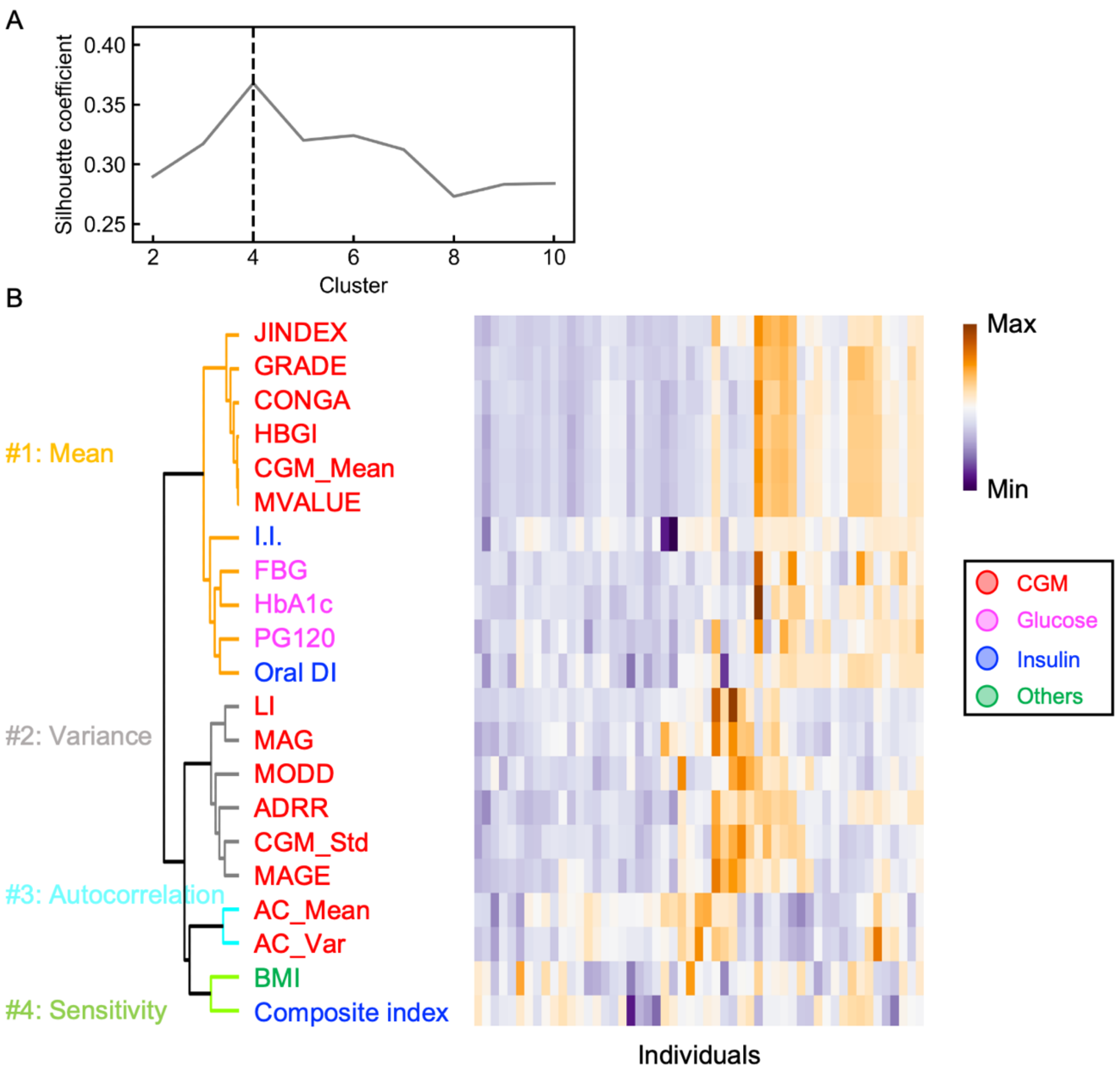


**Figure S4. Hierarchical clustering analysis of metabolic syndrome-related indices.**
(A) Relationship between the number of cluster and the silhouette coefficient. Dotted vertical line indicates the optimal number of clusters, which provides the best silhouette coefficient.
(B) Hierarchical clustering analysis of the standardized metabolic syndrome-related indices using Euclidean distance as a metric with the Ward method. The columns represent the standardized value of each index. The rows represent individuals. The indices are grouped and sorted according to their degree of relatedness.

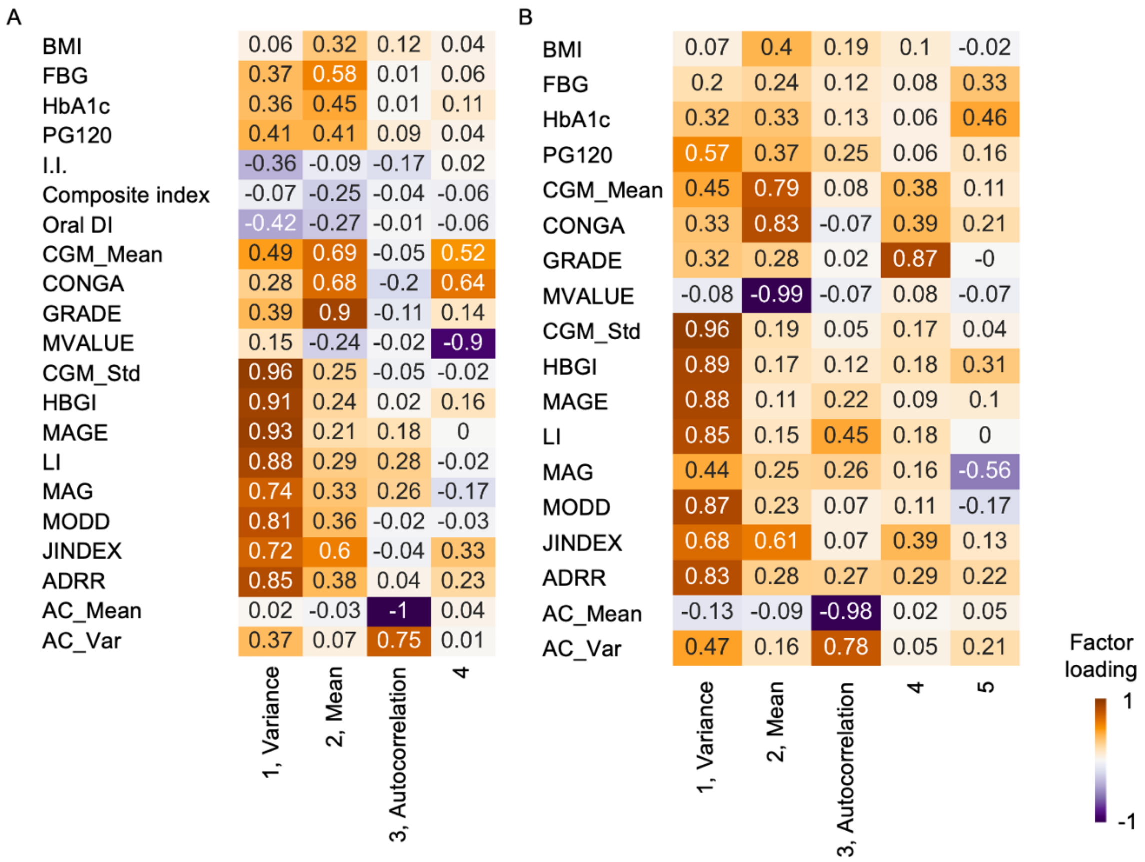


**Figure S5. Factor analyses of the clinical parameters using the previous datasets.** Factor analyses after orthogonal rotation. The values were based on the factor loadings. The columns represent each factor. The rows represent input variables. The analyses used the Japanese data from a previous study (Sugimoto et al., 2025) (A), and the American data from a previous study (Hall et al., 2018) (B).

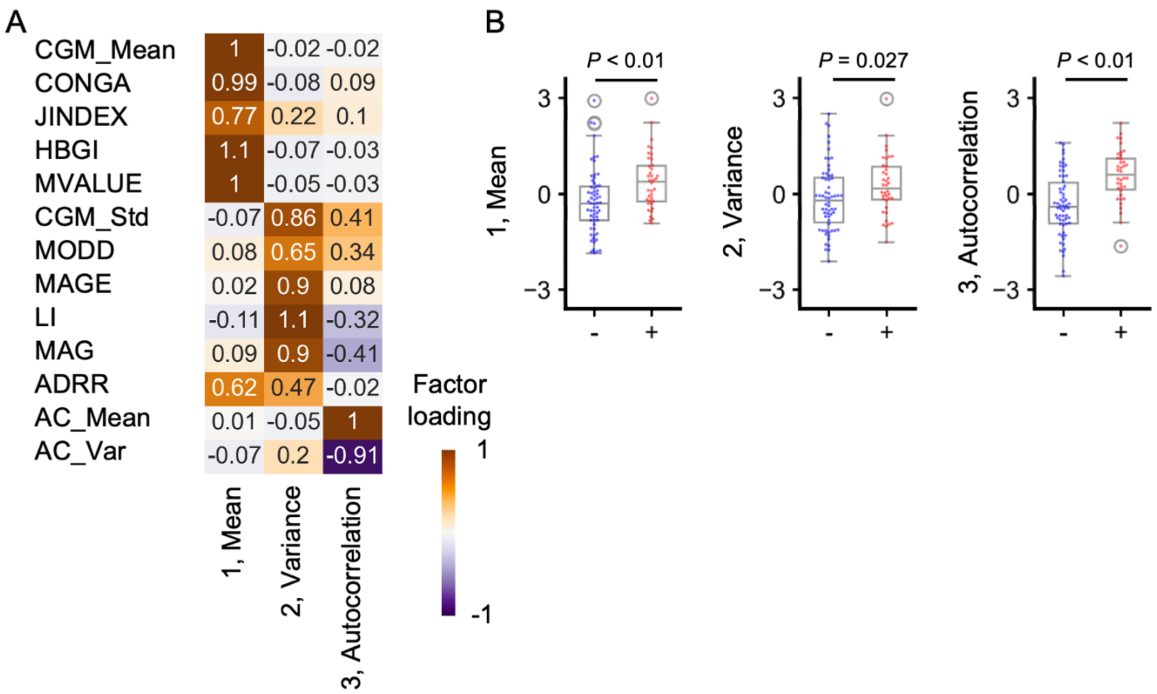


**Figure S6. Factor analysis of continuous glucose monitoring (CGM)-derived indices using a previously reported dataset.**

(A) Factor analysis of the CGM-derived indices. The heat map shows the factor loadings, with columns representing each factor and rows representing the input variables. The analysis used data from 100 Chinese individuals from a previous study (Zhao et al., 2023).

(B) Box plots comparing factors 1 (Mean), 2 (Variance), and 3 (Autocorrelation) between individuals without (-) and with (+) diabetic macrovascular complications. Each point corresponds to an individual. The boxes represent the interquartile range, with the median shown as a horizontal line. Mann–Whitney U tests were used to assess differences between groups, with $P$ values < 0.05 considered statistically significant.

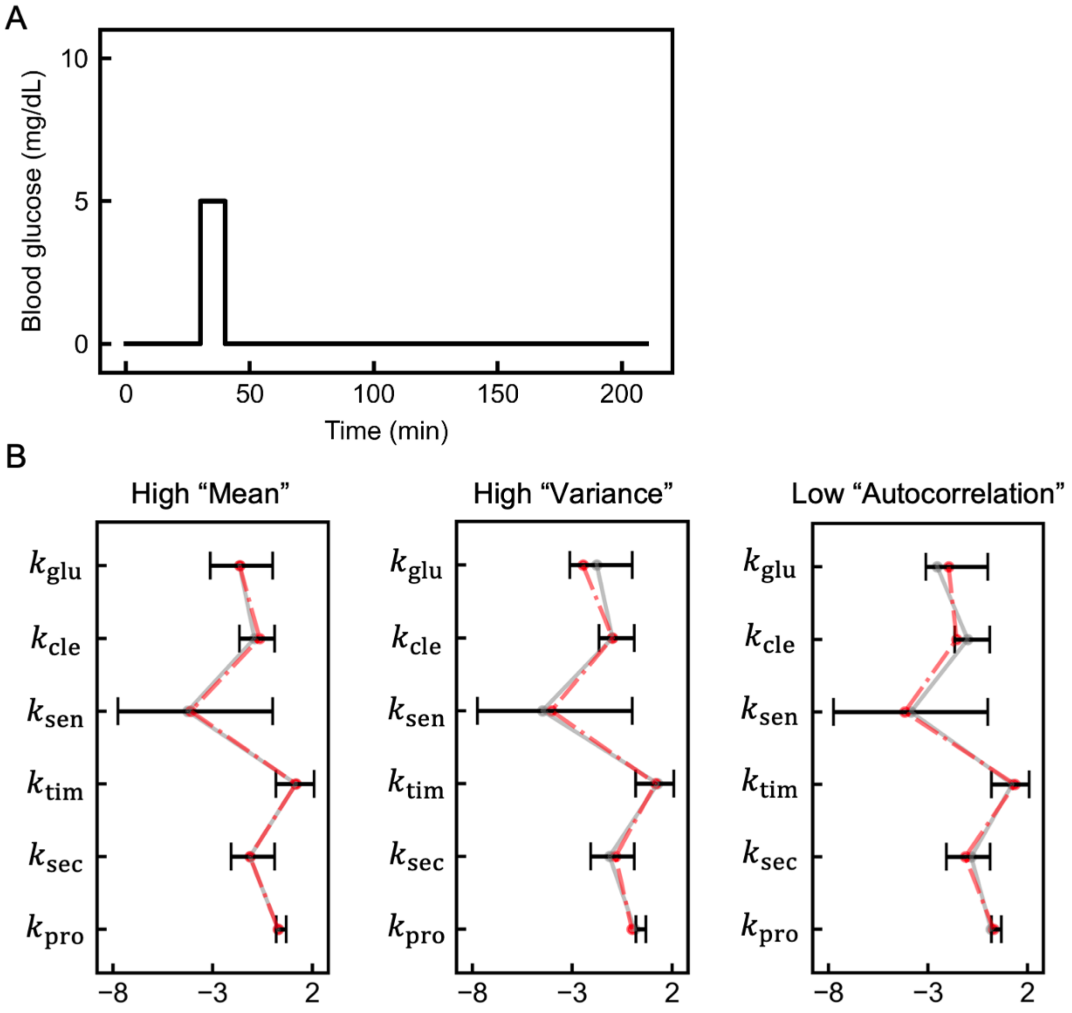


**Figure S7. Infused glucose and parameters used in simulating glucose dynamics.**
(A) The amount of the external input of glucose $f$ (see Methods).
(B) The parameters used in the simulation. The values of these parameters shown in red and gray correspond to the color of the simulated glucose dynamics shown in Figure 4A. Bars indicate the range of values for NGT individuals (De Gaetano and Arino, 2000).

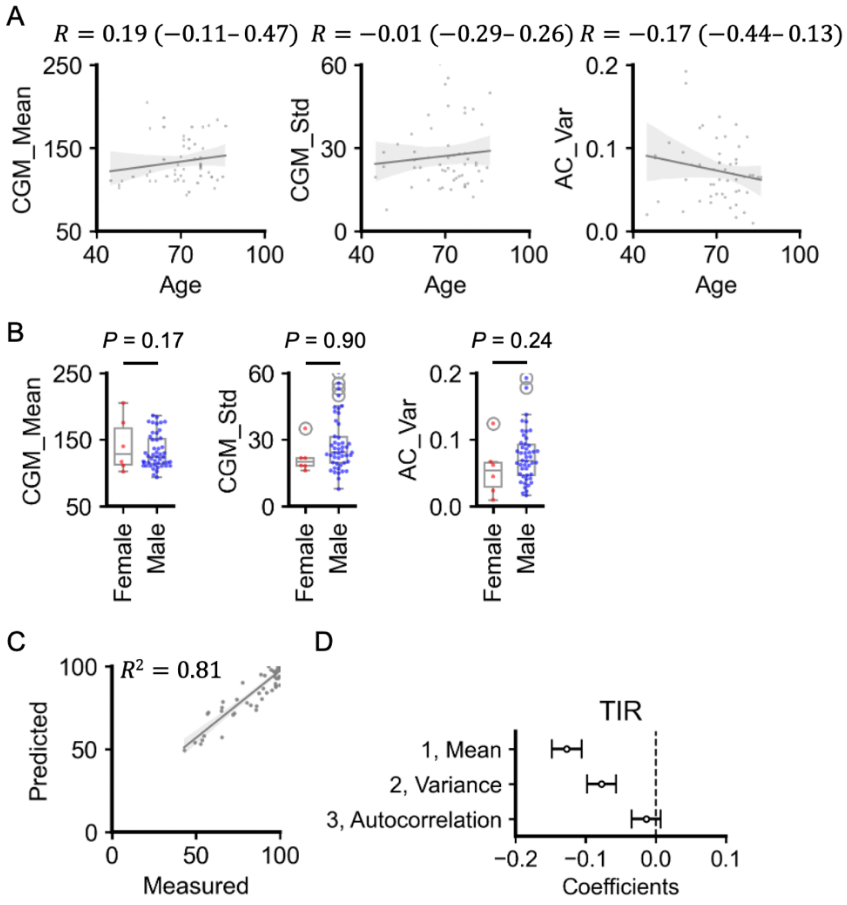


**Figure S8. Relation between CGM-derived measures and clinical measures.**

(A) Scatter plots and fitted linear regression lines for each CGM-derived measure versus age. Each point corresponds to the values for a single individual. Gray shaded area indicates the 95% CI.

(B) Box plots comparing CGM_Mean, CGM_Std, and AC_Var for females and males. Each point corresponds to an individual. The boxes represent the interquartile range, with the median shown as a horizontal line. Mann–Whitney U tests were used to assess differences between groups, with *P* values < 0.05 considered statistically significant.

(C) Comparison of predicted Time in range (TIR) versus measured TIR using multiple regression analysis between TIR and factor scores in Figure 3. In this analysis, TIR was the dependent variable, and the factor scores corresponding to the first three latent components (factor 1 representing the mean, factor 2 representing the variance, and factor 3 representing the autocorrelation) were the independent variables. Each point corresponds to the values for a single individual.

(D) Bars represent the 95% CIs of the coefficients of the regression model in (C).

**Table S1 Calculating formulae of the continuous glucose monitoring-derived indices.**

| Name | Formulae | Variables |
|---|---|---|
| CGM_Mean | $\frac{1}{N}\sum_{n=1}^{N} G_n$ | $G$ =glucose measured<br>$N$ =total number of readings |
| CGM_Std | $\sqrt{\frac{1}{N-1}\sum_{n=1}^{N}(G_n-\bar{G})}$ | $G$ =glucose measured<br>$N$ =total number of readings |
| CONGA | $\sqrt{\frac{1}{k-1}\sum_{t=t_1}^{t_k}(D_t-\bar{D})^2}$<br>$\bar{D}=\frac{1}{k}\sum_{t=t_1}^{t_k} D_t\,, D_t=G_t-G_{t-60}$ | $G$ =glucose measured<br>$k$ =number of observations with an observation 60 min ago<br>$t$ =time |
| LI | $\sum_{n=1}^{N-1}\frac{(G_n-G_{n+1})^2}{t_{n+1}-t_n}$ | $G$ =glucose measured<br>$N$ =total number of readings<br>$t$ =time |
| JINDEX | $0.324\times(\text{CGM\_Mean}+\text{CGM\_Std})^2$ | |
| HBGI | $\frac{1}{N}\sum_{i=1}^{N}\text{rh}(x_i)$ | $x$ = nonlinear transformation of glucose measured<br>$N$ =total number of readings<br>rh =risk value associated with a high glucose |
| GRADE | $\text{median}(425\times\{\log[\log(G_n)] + 0.16\}^2)$ | $G$ =glucose measured |
| MODD | $\frac{1}{k}\sum_{t=t_1}^{t_k}\lvert G_t-G_{t-1440}\rvert$ | $G$ =glucose measured<br>$k$ =number of observations with an observation 24 h ago<br>$t$ =time |
| MAGE | $\sum\frac{\lambda}{x}\quad if\ \ \lambda>v$ | $\lambda$ =blood glucose changes from peak to nadir<br>$x$ =number of observations<br>$v$ =1 Std of mean glucose for 24 h period |

| | | |
|---|---|---|
| ADRR | $$\frac{1}{N}\sum_{n=1}^{N}[\mathrm{LR}+\mathrm{HR}]$$ | $N$ =total number of readings<br>LR =risk value attributed to low glucose<br>HR =risk value attributed to high glucose |
| M-value | $$\frac{1}{N}\sum_{t=t_1}^{t_k}\left\|10\log\frac{18G_t}{\mathrm{IGV}}\right\|^3$$ | $G$ =glucose measured<br>$k$ = number of observations<br>IGV=ideal glucose value<br>$t$ =time |
| MAG | $$\frac{1}{T}\sum_{n=1}^{N-1}(G_n - G_{n+1})$$ | $G$ =glucose measured<br>$N$ =total number of readings<br>$T$ =total time |
| AC_Mean | $$\frac{1}{30}\sum_{l=1}^{30}\mathrm{AC}_l$$ | AC =autocorrelation of glucose<br>$l$ =lag |
| AC_Var | $$\frac{1}{29}\sum_{l=1}^{30}(\mathrm{AC}_l - \mathrm{AC\_Mean})^2$$ | AC =autocorrelation of glucose<br>$l$ =lag |
